# REM sleep microstructure alterations in REM sleep behavior disorder: beyond muscle tone

**DOI:** 10.1101/2025.01.02.24319683

**Authors:** Judith Nicolas, Louis Comperat, Patrice Fort, Anne Cheylus, François Ricordeau, Hélène Bastuji, Péter Simor, Laurène Leclair-Visonneau, Laure Peter-Derex

## Abstract

Isolated rapid eye movement (REM) sleep behavior disorder (iRBD) is characterized by dream enactment behaviors and loss of physiological atonia during REM sleep. It is considered a prodromal stage of alpha-synucleinopathies and may result from dysfunction of brainstem structures regulating muscle tone in REM sleep. Whether other REM sleep features are simultaneously affected remains unclear. Here, we investigated alterations in REM sleep microstructure, including phasic REM sleep, sawtooth waves (STW) and non-REM/REM sleep transitions, in iRBD and RBD associated with Parkinson’s disease (PD+RBD). We retrospectively included 20 patients with iRBD (85% male, 66.5[63–68]years), 20 patients with PD+RBD (75% male, 62.5[57.5-65]years) and 20 controls (75% male, 67[61–70]years). REM sleep without atonia (RSWA), bursts of REMs and STW bursts were manually scored. Phasic REM sleep proportion (derived from REMs), STW density/duration/frequency and the duration of NREM/REM transitions were compared between groups with a general linear mixed-effects model. Phasic REM sleep proportion was higher in the iRBD group (26.5[21–33]%) than in control (16.4[12.5-22.3]%, p-corrected=0.005) and PD+RBD (17.6[13.9-21.7]%, p-corrected=0.005) ones. Non-REM/REM transitions showed a duration gradient, increasing from controls (119.0[58.5-186.1]sec) to iRBD (212.1[68.5-391.4]sec, p-corrected=0.0038) and PD+RBD (375.8[217.6-514.6]sec, p-corrected<0.001) patients. STW density and duration were reduced in the PD+RBD group only (1.33[1.1-1.54]/min; 2.13[1.70-2.69]sec) vs controls (1.74[1.52-2.05]/min, p-corrected=0.005; 2.98[2.18-4.11], p-corrected<0.001), whereas altered STW spectral content was observed in both patient’s groups with a power shift toward higher frequencies (both p<0.001 vs controls). These results reinforce the hypothesis that REM sleep dysregulation in RBD extends to REM-specific electrophysiological features beyond loss of muscle atonia and dream enactments.

**Statement of Significance:** This study compared rapid eye movement (REM) sleep microstructure between patients with isolated REM sleep behavior disorder (iRBD) or RBD associated with Parkinson’s disease (PD+RBD), and controls. We found some altered REM sleep features in patient groups versus controls, as increased phasic REM sleep proportion in iRBD and reduced sawtooth waves (STW) density and duration in PD+RBD. Both groups exhibited altered STW spectral characteristics with a shift toward higher frequencies. Additionally, we observed a gradient in the duration of NREM/REM sleep transitions, increasing from controls to patients with iRBD and peaking in patients with PD+RBD, who exhibited the longest transitions. These findings highlight that REM sleep anomalies in RBD extends beyond atonia loss and dream enactments, encompassing broader microstructural changes.

## INTRODUCTION

Rapid eye movement (REM) sleep behavior disorder (RBD) is characterized by REM sleep without atonia (RSWA) which results in dream enactments with abnormal motor behaviors during REM sleep. Both of those diagnostic features, i.e. RSWA and motor behaviors, are currently assessed on video-polysomnography (v-PSG). RBD is caused by the dysfunction of REM sleep atonia-enabling structures that include the pontine subcoeruleus nucleus and the ventral medulla, as demonstrated by rodent model studies. ^[1,2]^ Damage to these structures in isolated RBD (iRBD) would be caused by the early accumulation of α-synuclein protein aggregates, whose progressive spread across the brain will eventually lead to overt α-synucleinopathies as Parkinson’s disease (PD), Lewy body dementia (LBD) or multiple system atrophy (MSA). ^[3]^ Other prodromal symptoms of α-synucleinopathies (such as dysautonomia, anosmia, cognitive impairment) exhibit a heterogeneous pattern of occurrence and progression in patients with iRBD, suggesting a patchy brain diffusion of pathological aggregates. ^[4]^ Given the various regions involved in REM sleep regulation and their widespread anatomical distribution, ^[5]^ it is likely that pathological damage in iRBD is not confined to structures generating REM sleep atonia. However, whether or not other structures involved in REM sleep expression are affected in iRBD remains unclear. Actually, few REM sleep features have been investigated using PSG beyond RSWA. Lower REM sleep stability, as assessed by stage shifts, was found in patients with iRBD and patients with PD compared to controls. ^[6]^ Regarding EEG spectral power, several works have shown the persistence of low frequencies during REM sleep, associated with cognitive impairment and/or a higher risk of conversion in α-synucleinopathy. ^[7–10]^ Results regarding faster frequencies are more contrasted: a decrease ^[11,12]^ or an increase ^[9,13]^ in beta-power were reported during (mostly phasic) REM sleep in patients with iRBD compared to controls. Such discrepancies might result from the heterogeneity of control groups, the topography of abnormalities, ^[13]^ and from the temporal dynamics of altered cortical activity. Indeed, the abnormal EEG slowing during REM sleep was recently reported to vanish over time, contrasting with a progressive increase in relative gamma power. ^[14]^ Besides, REM sleep encompasses several other microstructural features of interest than muscle atonia. ^[15]^

First, REM sleep can be divided into phasic and tonic REM sleep, two peculiar states defined respectively by the presence or absence of phasic events (such as rapid eye movements, REMs). ^[16]^ Of interest, these two micro-states have distinctive EEG pattern. More specifically, high (10-14 Hz) alpha and beta power is increased during tonic REM sleep, ^[17,18]^ which is considered to reflect a state close to relaxed wakefulness with a higher environmental alertness. ^[19,20]^ On the other hand, high frequency activity comprising gamma band is increased during phasic REM sleep, ^[18,21,22]^ which is considered to reflect intense internally generated sensorimotor, emotional and cognitive processes that may be linked to oniric experiences. ^[20,23–25]^ In iRBD, pathological motor behaviors predominantly occur during phasic REM sleep. ^[26,27]^ Yet, changes in phasic/tonic ratio have not been explicitly reported, although early reports have suggested an increase in REM density, ^[28,29]^ not confirmed later. ^[30–32]^ Interestingly, a recent study showed increased REMs density in RBD associated with PD, as compared to PD without RBD. ^[33]^

Second, EEG during REM sleep shows bursts of so-called sawtooth waves (STW) defined as trains of sharply contoured or triangular, often serrated, 2-5 Hz waves maximal in amplitude over the central head regions and often preceding a burst of REMs. ^[34,35]^ Their generators and functional role remain debated, yet, they are thought to be involved in synchronization of high frequency activities, as gamma band. ^[36,37]^ To our knowledge, although they represent a hallmark of REM sleep, STW waves have never been investigated in RBD context. According to previous studies, STW reduction can be observed in patients endowed thalamic or cortical damage. ^[38,39]^ Interestingly, STW have been suggested to represent the cortical component of PGO waves described in animal models and generated in the dorsal part of the nucleus subcoeruleus in the pons. ^[40–43]^ This suggests that brainstem lesions might also result in STW disruption.

Third, REM sleep microstructure features (muscle atonia, STW, bursts of REMs) gradually arise during the transition from NREM to REM sleep (NREM/REM sleep transition). This state alternation results from complex regulatory processes involving widespread brainstem and subcortical structures, mainly consisting of reciprocal inhibitory interactions between REM-off and REM-on neurons, ^[5]^ some of which are thought to be impaired in iRBD, as well as by top-down processes of regulation involving cortical areas such as the medial prefrontal cortex. ^[44]^ Hence, NREM/REM sleep transitions are all the more interesting in RBD, since they have been shown to be lengthened in patients with brainstem damage in the context of bulbar post-polio syndrome. ^[45]^

Investigating all these REM sleep features in iRBD could lead to a better understanding of the extent of REM sleep dysfunction, which may go beyond motor disturbances as recently suggested in the context of RBD associated with PD. ^[46]^ In addition, a gradient of impairment in these features between iRBD and RBD associated with PD would suggest that they could provide promising biomarkers of phenoconversion, since all available as early as diagnostic v-PSG is performed. In this study, we thus explored the proportion of phasic REM sleep, the characteristics of STW and the duration of NREM/REM sleep transitions in iRBD patients, patients with PD and RBD compared to controls. We hypothesized that in patients with RBD compared to controls, (i) the proportion of phasic REM sleep would be increased, (ii) STW density and amplitude would be decreased, with altered spectral signatures, and (iii) NREM/REM sleep transitions would be longer. Such alterations would be more pronounced in patients with PD and RBD compared to patients with iRBD, as a result of more widespread lesions in structures regulating REM sleep, especially for features exhibiting a cortical component such as STW.

## METHODS

### Participants

For inclusion, we retrospectively screened 56 patients with iRBD who benefited from a v-PSG recording in the Center for Sleep Medicine, University Hospital of Lyon, between 2019 and 2023. Exclusion criteria were: medications that may influence REM sleep (as antidepressants, anxiolytics, neuroleptics and betablockers) and moderate or severe obstructive sleep apnea syndrome with an apnea-hypopnea index (AHI) above 20/h (the usual threshold of 15/h usually retained for defining moderate or severe OSA was extended to 20/h given the age of the population included and the high prevalence of respiratory abnormalities even in non-clinical populations). ^[47]^ Among iRBD patients screened, 20 met the inclusion criteria (17 with AHI > 20/h, 12 with REM sleep-interfering treatment and 7 were excluded based on both criteria) and were included in the study (iRBD group; 17/20 i.e. 85% of males; median [Q1-Q3] age: 66.5 ^[63–68]^ years; median reported RBD symptoms duration at the time of the v-PSG: 5.9 years [3.5-8.3]). Twenty control individuals matching for age, sex and AHI with the iRBD group were selected among patients free of any neurological disease who underwent a PSG in the same Center as part of routine obstructive sleep apnea screening (control group; 15/20 i.e. 75% of males; age: 67 ^[61–70]^ years). Finally, 20 age-and sex-matched patients with PD and RBD were selected from a research cohort of the Sleep Center of the University Hospital of Nantes (PD+RBD group; 15/20 i.e. 75% of males; age: 62.5 [57.5-65] years; median PD duration: 8.8 years [5.9-11.5]). The medication exclusion criterion was not applied to this latter group because most of these patients were taking psychotropic drugs, especially as their dopaminergic treatment was not discontinued before the v-PSG recording. For iRBD and PD+RBD groups, RBD was diagnosed based on clinical data and a v-PSG, according to the ICSD3-TR criteria. ^[48]^

The study was performed in accordance with the principles of good clinical practice and the Declaration of Helsinki. The trial was approved by the Ethic Review Board of the Hospices Civils de Lyon (N° 23-5452 issued on 21/03/2024). All participants provided informed consent.

### PSG recordings

All subjects underwent a full-night in-hospital PSG, with video for patients with RBD. PSG were conducted with Natus® (3 iRBD, 18 controls and 20 PD+RBD) or Micromed® (17 iRBD and 2 controls) acquisition system. The following signals were recorded (sample rate: 256 Hz): electroencephalography (EEG), electrooculography (EOG), electromyography (EMG) of the chin and tibialis muscles, electrocardiography, pulse oximetry by finger probe, nasal pressure transducer and naso-buccal thermistor, chest and abdominal respiratory movements recording, and microphone. EEG montages could vary depending on suspected diagnoses and on the center where PSG recordings were performed (positioning of electrodes according to the International 10-20 System): Fp2, C4, T4, O2 in all patients, and (1) Fp1, C3, T3, O1, Fz, Cz, Pz in the iRBD group, (2) Fz, Cz, Pz in the control group, (3) Fp1, C3, O1, T3 in the PD+RBD group.

### Signal analysis

#### Visual analysis of REM sleep microstructure and NREM/REM sleep transitions

EEG data were band passed filtered between 0.3 and 70 Hz, EOG data between 0.3 and 15 Hz and EMG data between 10 and 200 Hz. Sleep stages and sleep-related events were scored according to the AASM guidelines. ^[49]^

REM sleep macrostructure was characterized by the REM sleep duration, the REM sleep percentage (with respect to the total sleep time), and the REM sleep latency. REM sleep stability was also assessed by the arousal index in REM sleep as well as the number, index (number of REM bouts divided by the time spent in REM sleep) and duration of REM sleep bouts.

Then, the following features were visually double-marked by two neurologists (L.C. and L.P.-D.): STW, bursts of REMs, onset of chin muscle atonia and RSWA. These features were marked during all REM sleep bouts and during the 10 minutes preceding the onset of a REM sleep episode, according to a recent study indicating that STW appeared on average 4 minutes before the first epoch scored as REM sleep, ^[50]^ thus suggesting that NREM/REM sleep transitions could start several minutes before actual REM sleep scoring.

STW were defined as bursts of consecutive surface-positive 2-5Hz frontocentral bilateral synchronous symmetric waves with an amplitude of 20-100 mV, an angle >80°, and with a slow incline to a negative peak with a following steep linear decline ending in a positive peak. ^[34]^ STW segments median duration was computed for each subject and STW density was calculated as the number of STW segments per minute of REM sleep.

REMs were detected on the EOG as steep-front deflections lasting less than 500 ms and out of phase on the two EOG channels (corresponding to a combined movement of both eyes). ^[49]^ REMs periods were marked by segments of at least 3 seconds, and periods separated by less than 3 seconds were grouped into a single REMs period. ^[36]^ Phasic and tonic REM sleep corresponded to periods with and without REMs respectively. The phasic REM sleep ratio was defined as the phasic REM sleep duration with respect to the total REM sleep duration. To investigate the evolution of the phasic ratio across the night, the total sleep period was divided in four equal time bins. The specific phasic ratio of each night quarter was computed and normalized within each group with the mean and the standard deviation of the four night periods.

Muscle atonia onset was determined by the absence of chin EMG signal for at least 1 second and was marked once for each NREM/REM sleep transition. RSWA percentage was calculated as the number of 3-second mini-epochs containing “any” chin EMG activity (i.e. EMG activity > 2 times the background EMG lasting > 0.1 sec) divided by the total number of 3-second mini-epochs during REM sleep after exclusion of arousals, according to the SINBAR (Sleep Innsbruck Barcelona) scoring method validated by the International RBD Study Group. ^[51,52]^ We chose to quantify RSWA on the chin EMG only, for homogenization purposes, as only a subset of patients had flexor digitorum superficialis EMG, as recommended. ^[51]^ RSWA threshold was set to 14% as specificity for iRBD diagnosis has been reported to be maximal when using this quantification method. ^[53]^

Finally, we investigated the sequence of the three events (muscle atonia onset, first STW, first REMs) during the NREM/REM sleep transitions and duration of these transitions. The sequence of events was analyzed as a categorical variable of 6 possible transition sequences. The duration of NREM/REM sleep transitions was defined, for each REM sleep episode, as the absolute difference between the first STW burst and the first burst of REMs. Indeed, several studies have reported STW to precede the onset of REMs during NREM/REM transitions ^[34,50]^. We chose to exclude atonia onset from the definition of these transitions, as it was biased by the presence of muscle tone abnormalities in RBD.

#### STW spectral analysis

For the EEG spectral analysis, data were further visually screened with the aforementioned filters to reject segments contaminated with artifacts. Investigated channels were selected to be common to the three groups of participants (F2, C4, and O2). Using mne python (version 1.7.1), C4 was used to re-reference signal on F2 and O2 electrodes. Then, an independent component analysis was applied on high-passed filtered (0.1 Hz) artifact-free data to remove physiological noise such as eye movements. STW segments occurring during REM sleep were extracted and the power spectral density (PSD) of each STW segment was estimated with the Welch method (Hamming window, 1 sec duration, no overlap). The FOOOF algorithm (version 1.0.0)^[54]^ was used to parameterize neural power spectra on F2-C4 and O2-C4 separately. Settings for the algorithm were set as follows 1) peak width limits: 0.5 and 12.0; 2) max number of peaks: 4; 3) minimum peak height: 0; 4) peak threshold: 2; and 5) aperiodic mode: fixed. Power spectra were parameterized across the frequency range 0.5 to 10 Hz. STW clean segments of more than 2 seconds were further analyzed. For each segment, we extracted the following parameters of the channel-specific fits: time of the segment, segment duration, segment order in the night, and peak order in the fit. Note that peaks were sorted by height (over and above the aperiodic component). For each detected peak of individual segments, we extracted the center frequency, the power, and the bandwidth.

### Statistics

Statistical analyses were performed with R software (version 4.4.1). ^[55]^ Continuous quantitative variables are presented as medians and interquartile ranges (IQR), and qualitative variables are presented as N (%).

Qualitative variables, such as the sequence of events at NREM/REM sleep transitions, were analyzed for an overall comparison using Fisher’s exact test and the post hoc pairwise comparison using FDR correction.

Dependent quantitative variables evaluating REM sleep structure and features (REM sleep macro and microstructure parameters, RSWA, proportion of phasic REM sleep and STW index and duration) were compared using a general linear mixed-effects model (LMM) with the group factor (control vs. iRBD vs. PD+RBD) as a fixed effect and the participant as random effect. For STW spectral characteristics, we first explored the frequency profile of STW segments occurring during REM sleep. To do so, we determined in the frequency distribution the mode of the maximum peak of each segment regardless the channel, the group and the participant. We found that the frequency distribution of the oscillatory activities in the STW segments was bimodal: (i) The low frequency distribution was characterized by a mode of 2.50 Hz, a kernel density estimates at the mode of 0.32 and a proportion of area underneath the mode of 0.39; (ii) The high frequency distribution was characterized by a mode of 4.37 Hz, a kernel density estimates at the mode of 0.26 and a proportion of area underneath the mode of 0.61. We thus decided to divide the fitted peaks according to two frequency profiles: low or high frequency, which were defined according to a frequency limit computed as the mean between the low and the high frequency modes. Then, we extracted the number of segments according to the frequency profile of the maximal fitted peak, for each participant and each channel. A general linear mixed models with random effects (participant) was developed to analyze the number of segments according to their frequency profile (low vs. high) in the three populations (control vs. iRBD vs. PD+RBD) and the two channels (F2 vs. O2). We also interrogated the power of the fitted peaks with a general linear mixed model with random effects (participant, peak number in the fit, STW segment time of the night and STW segment duration) developed to analyze the number of peaks according to their frequency profile (low vs. high) in the three populations (control vs. iRBD vs. PD+RBD) and the main effect of channels (F2 vs. O2).

Normality and homogeneity of the residuals of the LMM were assessed visually using Visual LMM assumptions (normality of residuals, normality of random effects, linear relationship, homogeneity of variance, multicollinearity). If the assumptions were not met, data transformations were applied (i.e., square-root or log transformations). Type-III ANOVAs were executed to extract significant effects (F statistics and eta squared (η^2^) effect size are reported). For LMM post-hoc comparisons, FDR corrections were applied. When data transformation was not sufficient to meet the assumptions, ANOVA which uses permutation tests instead of normal theory tests (partial η^2^ effect size are reported) and pairwise Wilcoxon tests were used for post-hoc comparisons (W statistics and r effect size are reported). False Discovery Rate (FDR) corrections were applied for significant effects of the statistical models for all post hoc analysis and only the corrected p-values are presented.

## RESULTS

Demographic data and PSG variables retrieved in the three groups are summarized in Table 1.

**Table 1.** Demographic and polysomnography characteristics provided as median [IQR] across participants separately for each group. PSG: polysomnography; TST: total sleep time; WASO: wake after sleep onset; REM: rapid eye movement; RBD: REM sleep behavior disorder; iRBD: isolated RBD; PD: Parkinson’s disease.

| Variable, | Controls, n = 20 | iRBD, n = 20 | PD+RBD, n = 20 |
| --- | --- | --- | --- |
| Age, years | 67 [61-70] | 66.5 [63-68] | 62.5 [57.5-65] |
| Sex, male | 15 (75) | 17 (85) | 15 (75) |
| PSG measures |  |  |  |
| TST, min | 388 [345-446] | 458 [415-496] | 390 [338-426] |
| Sleep cycles | 4 [4-5] | 4 [3.75-5] | 3.5 [2.75-4] |
| Sleep efficiency, % | 76.2 [70.8-82.2] | 81.1 [73.1-86] | 65 [61-76.2] |
| WASO, min | 89.5 [47.5-108] | 81 [56-114] | 89 [64-128] |
| Stage N1, % | 6.15 [3.9-12.3] | 5.15 [3.65-8.8] | 11.2 [8.97-18.5] |
| Stage N2, % | 50 [40.8-58.3] | 49.1 [42.5-55.1] | 48.2 [43.9-59.7] |
| Stage N3, % | 19.4 [12.1-28.1] | 21.5 [18.4-24.2] | 18.4 [14.2-26.1] |
| Stage REM, % | 21.6 [17.3-26.3] | 19.1 [13.3-24.6] | 13.8 [10.9-19] |
| Arousal index, /h | 14.7 [12.1-17] | 22.7 [14.9-26.9] | 15.6 [11.3-17.3] |

### REM sleep macrostructure

REM sleep macrostructure variables are reported in Table 2. The LMM on REM sleep duration (min) and proportion (% of total sleep time) revealed a significant main effect of group (duration: F(2,57) = 6.25, p = 0.0035, η^2^ = 0.18; percentage: F(2,57) = 5.71, p = 0.0055, η =0.17). Post-hoc analysis revealed that both variables were significantly lower in the PD+RBD group (duration: 52.5 min [36.0 – 69.5], 13.8 % [10.9 – 19.0]) compared to controls (duration: 87.2 min [61.8 – 100.0], t(57) = 2.56, p = 0.02; percentage: 21.6 % [17.3 – 26.3], t(57) = 3.31, p = 0.0049) and iRBD (duration: 84.7 min [69.4 – 106.0], t(57) = 3.39, p = 0.0038; percentage: 19.1 % [13.3 – 24.6], t(57) = 2.24, p = 0.044) groups. REM sleep duration and percentage was not different between controls and iRBD (duration: t(57) = -0.84, p = 0.41; percentage: t(57) = 1.08, p = 0.29).

**Table 2.**
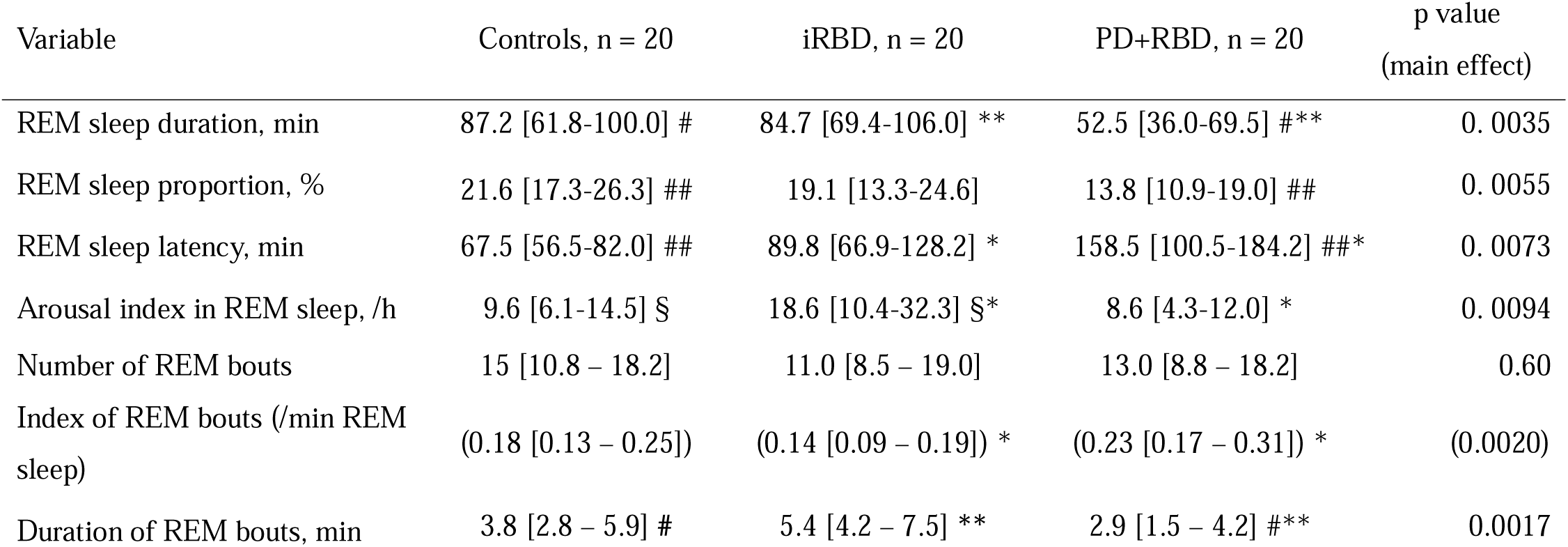
REM sleep macrostructure. REM sleep macrostructure characteristics are provided as median [IQR] in each group of participants. Post-hoc analysis using Tukey HSD consisted in three comparisons and are reported if significant: **#** refers to a p-value < 0.050 when comparing PD+RBD and controls (##: p < 0.010); **§** refers to a p-value < 0.050 when comparing iRBD and controls; **\*** refers to a p-value < 0.050 when comparing PD+RBD and iRBD (**: p < 0.010). REM, rapid eye movement; RBD, REM sleep behavior disorder; iRBD, isolated RBD; PD, Parkinson’s disease.

The LMM on REM sleep latency revealed a significant main effect of group (F(2,57) = 5.37, p = 0.0073, η^2^ = 0.16). Post-hoc analysis revealed that REM sleep latency was longer in the PD+RBD group (158.5 min [100.5 – 184.2]) compared to the two other groups (controls: 67.5 min [56.5 – 82.0], t(57) = -3.09, p = 0.0094; iRBD: 89.8 min [66.9 – 128.2], t(57) = -2.50, p = 0.023). REM sleep latency was not different between controls and iRBD patients (t(57) = - 0.59, p = 0.56).

The LMM on the arousal index during REM sleep revealed a significant main effect of group (F(2,57) = 5.07, p = 0.0094, η^2^ = 0.15). Post-hoc analysis revealed that arousal index during REM sleep was higher in the iRBD patients (18.6/h [10.4 – 32.3]) compared to controls (9.6/h [6.1 – 14.5], t(57) = -2.48, p = 0.024) and to PD+RBD patients (8.6/h [4.32 – 12], t(57) = 2.97, p = 0.013). The arousal index during REM sleep did not differ between controls and PD+RBD patients (t(57) = 0.49, p = 0.63).

The LMM on REM bouts number did not reveal a significant effect of group (F(2,57) = 0.52, p = 0.60, η^2^ = 0.018) whereas the REM bouts index did (F(2,57) = 6.97, p = 0.0020, η^2^ = 0.20). Post-hoc analyses revealed that the normalized number of REM bouts was higher in the PD+RBD (0.23 [0.17 – 031]) than in the iRBD group (0.14 [0.009 – 0.19], t(29.86) = -3.46, p = 0.002 (0.005 FDR-corrected), Cohen’s d = -1.09). The normalized number of REM bouts of control participants (0.18 [0.13 – 0.25]) did not significantly differ from iRBD (t(37.41) = 1.81, p = 0.078 (0.078 FDR-corrected), Cohen’s d = 0.57) nor from PD+RBD patients (t(32.20) = -2.05, p = 0.049 (0.074 FDR-corrected), Cohen’s d = -0.65).

The LMM assessing REM bout durations met the required assumptions after data log transformation. This analysis revealed a group main effect (F(2,47.89) = 7.32, p = 0.0017, η^2^ = 0.23). REM bout durations were lower in the PD+RBD group (2.9 min [1.5 – 4.2]) as compared to the control (3.8 [2.8 – 5.9], t(48.5) = 2.64, p = 0.011 (0.017 FDR-corrected)) and the iRBD groups (5.4 [4.2 – 7.5], t(52.7 = 3.52, p < 0.001 (0.0027 FDR-corrected)). These durations did not differ between control and iRBD participants (t(48.5) = 1.09, p = 0.28 (0.28 FDR-corrected)).

### REM sleep without atonia

The LMM assessing the RSWA percentage did not meet the required assumptions even after data transformation. We thus performed an ANOVA with permutation test, with group as fixed effect and participant as random effect. This analysis revealed a group main effect (Figure 1, p < 0.001, η^2^ = 0.64). Post-hoc analyses revealed that RSWA percentage was higher in the iRBD group (62.2 % [46.6 – 72.9]) as compared to controls (3.7 % [2.4 – 6.1], W = 0.00, p < 0.001 (< 0.001 FDR-corrected), r = 0.85) and PD+RBD (28.5 % [20.1 – 51.8], W = 318, p = 0.001 (0.002 FDR-corrected), r = 0.49). RSWA percentage was higher in the PD+RBD group as compared to controls (W = 2, p < 0.001 (< 0.001 FDR-corrected), r = 0.85).

**Figure 1.**
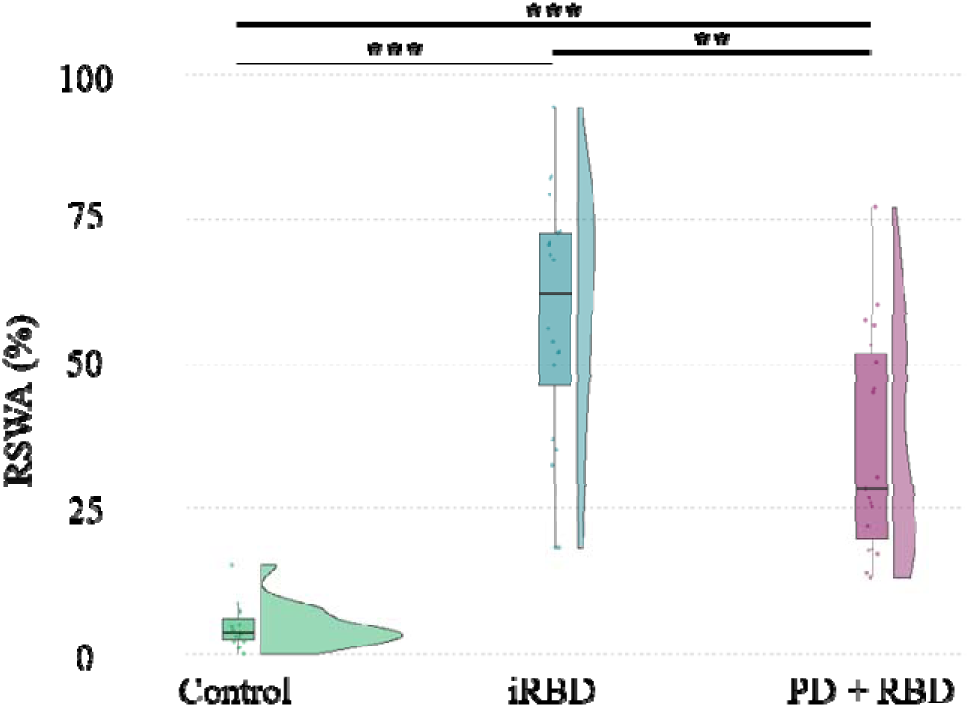
REM Sleep Without Atonia (RSWA) percentage. RSWA percentage is higher in the iRBD than in the PD+RBD and in the control groups. Statistics are displayed for each group with a density plot (kernel density estimate) and a boxplot: the bold line corresponds to the median; lower and upper hinges correspond to first and third quartiles; lower and upper whiskers extend from the lower or upper hinges to the lowest or largest value no further than 1.5xIQR from the hinge. Colored data points correspond to participant’s individual values, which are jittered in arbitrary distances on the x-axis within the respective boxplot to increase perceptibility. **: p<0.01; ***: p<0.001; n.s.: non-significant. RBD, rapid-eye movement sleep behavior disorder; iRBD, isolated RBD; PD, Parkinson’s disease; RSWA, REM sleep without atonia; EMG, electromyography; IQR, interquartile range.

### Phasic REM

The LMM assessing the phasic REM sleep ratio (defined as the ratio of the phasic REM sleep duration to the total REM sleep duration) did not meet the required assumptions even after data transformation. We thus performed an ANOVA with permutation test with group as fixed effect and participant as random effect. The permutation ANOVA revealed a significant main effect of group (Figure 2A, p < 0.001, η^2^ = 0.22). Phasic REM sleep ratio was significantly higher in the iRBD group (26.5 % [21.0 – 33.0]) than in the controls group (16.4 % [12.5 – 22.3], W = 94, p = 0.004 (0.005 FDR-corrected), r = 0.45) and in the PD+RBD group (17.6 % [13.9 – 21.7], W = 308, p = 0.003 (0.005 FDR-corrected), r = 0.46). However, there was no statistically significant difference between PD+RBD and controls (W = 194, p = 0.88 (0.88 FDR-corrected), r = 0.026).

**Figure 2.**
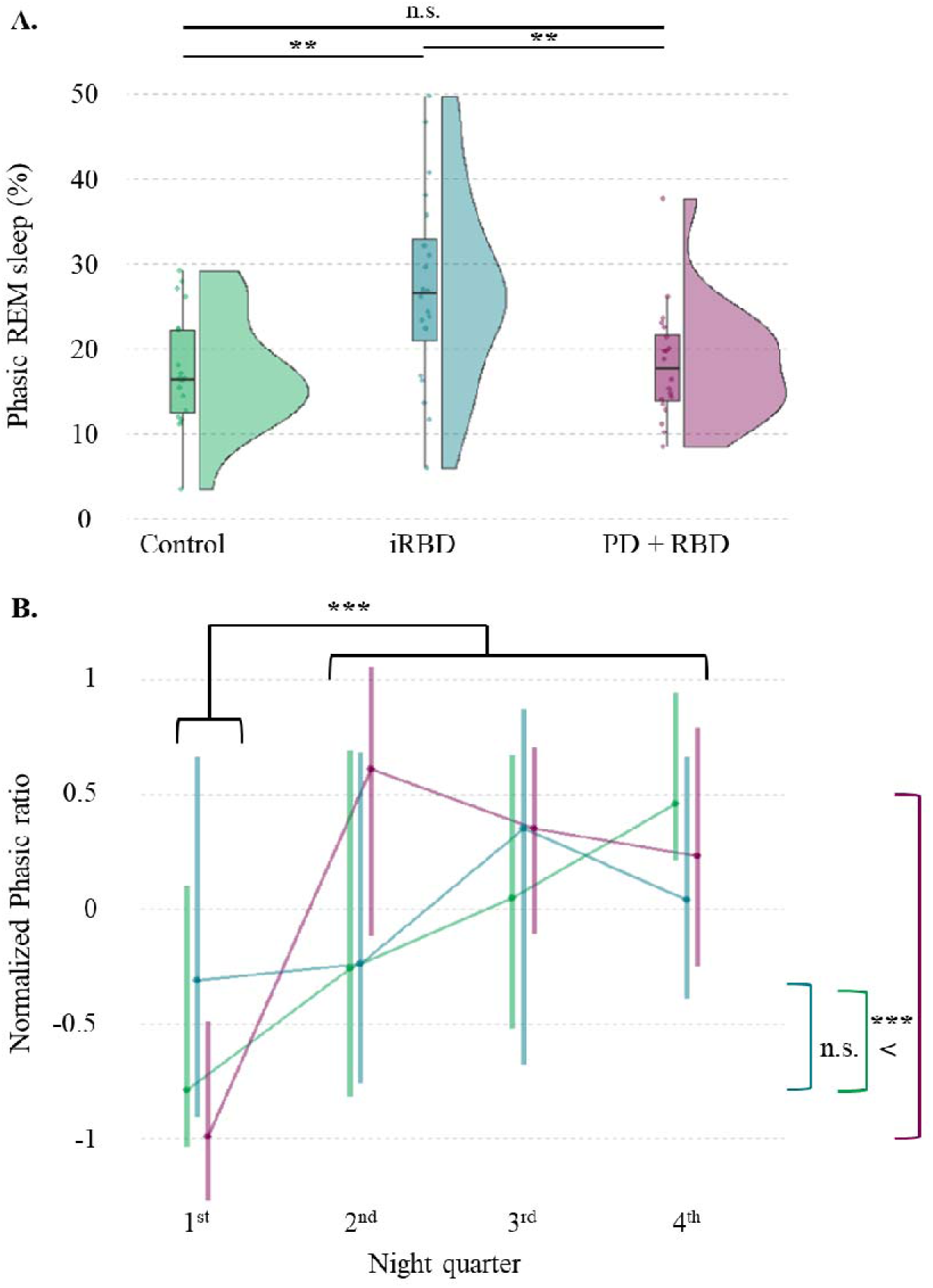
Phasic REM sleep percentage of total REM sleep across controls, patients with iRBD and patients with PD and RBD. A. Phasic REM sleep percentage is significantly higher in the iRBD than in the control and the PD+RBD groups. Statistics are displayed for each group with a density plot (kernel density estimate) and a boxplot: the bold line corresponds to the median; lower and upper hinges correspond to first and third quartiles; lower and upper whiskers extend from the lower or upper hinges to the lowest or largest value no further than 1.5xIQR from the hinge. Colored data points correspond to participant’s individual values, which are jittered in arbitrary distances on the x-axis within the respective boxplot to increase perceptibility. B. The normalized phasic ratio increases across night quarters. The increase from the first to the other night quarters is bigger in the PD + RBD group. Statistics are displayed for each group with the median and the IQR, which are jittered in arbitrary distances on the x-axis within the respective boxplot to increase perceptibility. **: p<0.01; n.s.: non-significant. REM: rapid eye movement; RBD: REM sleep behavior disorder; iRBD: isolated RBD; PD: Parkinson’s disease; IQR: interquartile range

To investigate phasic REM sleep regulation across the night, we explored the phasic ratio in each night quarter (Figure 2B). First, phasic ratio was normalized across night quarter to account for differences between groups in overall phasic ratio and simplify the LMM which only assessed night quarter main effect and its interaction with the group. The LMM met the required assumptions without transformation. This analysis revealed a main effect of the night quarter (F(3,228) = 10.12, p < 0.001, η^2^ = 0.63). The interaction between the group and the night quarter was also significant (F(6,228) = 2.96, p = 0.0084, η^2^ = 0.37). Post analyses revealed that the normalized phasic ratio was lower for the first night quarter than for the others (vs. 2^nd^: t(228) = -3.80, p < 0.001, Cohen’s d = -0.63; vs 3^rd^ : t(228) = -4.31, p < 0.001, Cohen’s d = -0.74; vs 4^th^: t(228) = -5.04, p < 0.001, Cohen’s d = -0.95). The other night quarters did not show differences from one another (2^nd^ vs 3^rd^: t(228) = -0.51, p = 0.61, Cohen’s d = -0.87; 2 vs. 4^th^: t(228) = -1.24, p = 0.33, Cohen’s d = -0.23; 3^rd^ vs. 4^th^: t(228) = - 0.73, p =0.56, Cohen’s d = ²-0.14). We thus restricted the post hoc analysis on the group x night quarter interaction to the difference between the first and the other quarters. We found that the increase in phasic ratio from the 1^st^ to the 2^nd^ quarter was significantly greater in the PD + RBD group than in the control (t(171) = 2.52, p = 0.013 (0.041 FDR-corrected)) and the iRBD group (t(171) = 3.61, p < 0.001 (0.0036 FDR-corrected)). The difference between the 1^st^ and the 3^rd^ and the 1^st^ and the 4^th^ remained significant when comparing the PD + RBD and the iRBD group (t(171) = 2.50, p = 0.014 (0.041 FDR-corrected); t(171) = 2.38, p =..019 (0.042 FDR-corrected), respectively).

### STW analysis

A total of 7455 STW segments were marked during REM sleep (controls: 3052, iRBD: 2811, PD+RBD: 1592). The LMM assessing the STW density during REM sleep did not meet the required assumptions including after data transformation. We thus performed an ANOVA with permutation test with group as fixed effect and participant as random effect. The permutation ANOVA revealed a significant main effect of group (Figure 3A, p = 0.043, η^2^ = 0.16). STW density was significantly lower in the PD+RBD group (1.33/min [1.1 – 1.54]) compared to the controls (1.74/min [1.52 – 2.05], W = 313, p = 0.002 (0.005 FDR-corrected), r = 0.48). STW density was not significantly different between the iRBD group (1.47/min [1.18 – 1.91]) and both other groups (vs. control: W = 240, p = 0.29 (0.29 FDR-corrected), r = 0.17; vs. PD+RBD: W = 257, p = 0.13 (0.19 FDR-corrected), r = 0.24).

**Figure 3.**
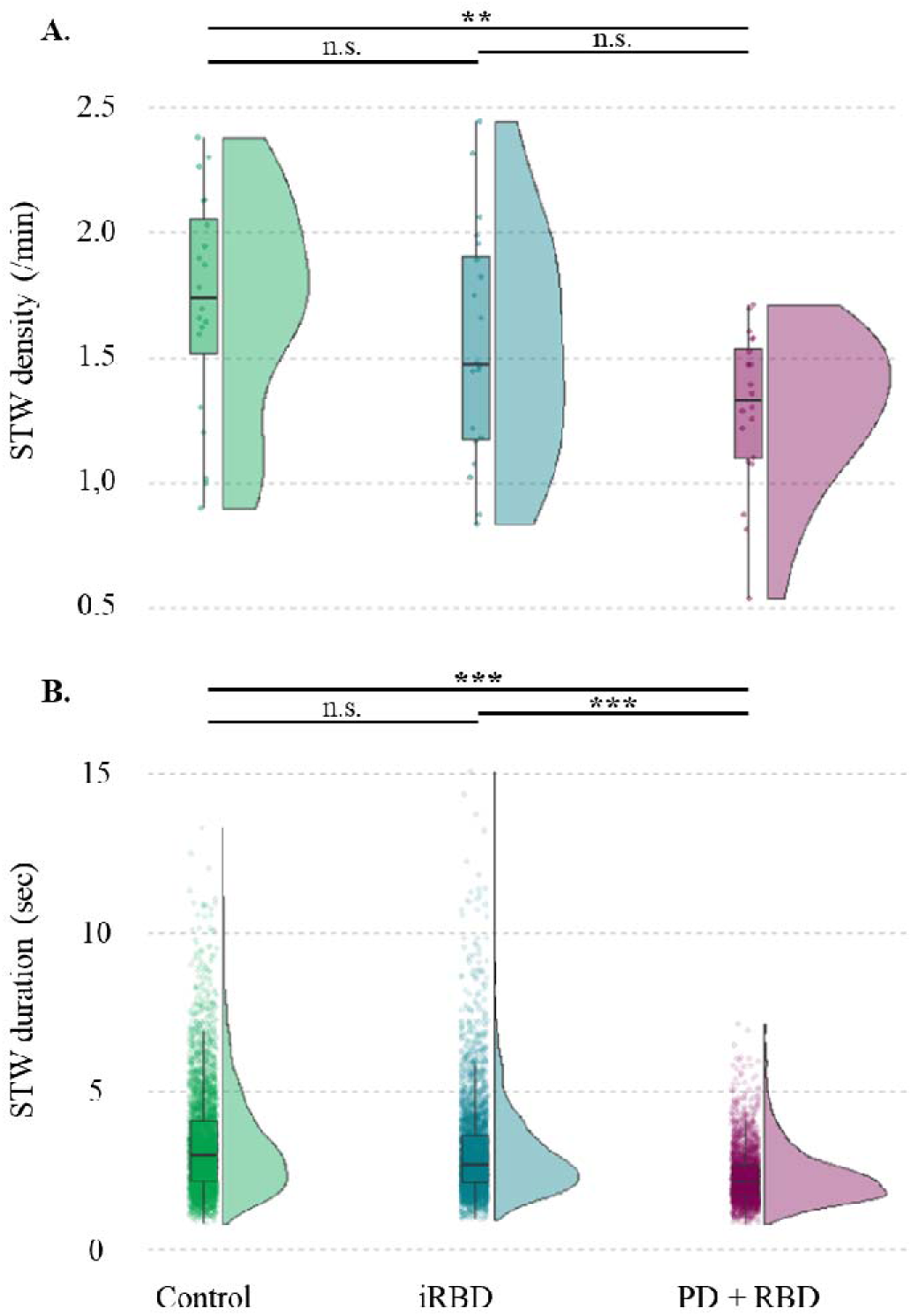
Sawtooth waves (STW) characteristics. STW density (A.) is lower in the PD+RBD than in the controls group. STW segments duration (B.) is shorter in the PD+RBD than in the control and the iRBD groups. Statistics are displayed for each group with a density plot (kernel density estimate) and a boxplot: the bold line corresponds to the median; lower and upper hinges correspond to first and third quartiles; lower and upper whiskers extend from the lower or upper hinges to the lowest or largest value no further than 1.5xIQR from the hinge. Colored data points correspond to participant’s individual values (A) or to individual STW segments (B.), which are jittered in arbitrary distances on the x-axis within the respective boxplot to increase perceptibility. **: p<0.01; ***: p<0.001; n.s.: non-significant. STW: sawtooth waves; RBD: rapid-eye movement sleep behavior disorder; iRBD: isolated RBD; PD: Parkinson’s disease; IQR: interquartile range.

The LMM assessing the STW duration during REM sleep in function of the groups met the required assumptions after log transformation of the data. The ANOVA revealed a significant main effect of group (Figure 3A, F(2,56.67) = 21.61, p < 0.001, η^2^ = 0.41). Post-hoc analyses revealed that STW segment duration was significantly lower in the PD+RBD group (Figure 3B, 2.13 sec [1.70 – 2.69]) compared to the controls (2.98 sec [2.18 – 4.11], z = 5.58, p < 0.001) and to the iRBD (2.71 sec [2.13 – 3.59], z = 5.68, p < 0.001) but did not differ between controls and iRBD (z = -0.11, p = 0.91).

Spectral analysis performed on 5153 STW segment > 2 sec sleep (controls: 2254, iRBD:2095, PD+RBD: 804) showed a peak around 2.5 Hz and a second peak around 4.5 Hz (Figure 4A).

**Figure 4.**
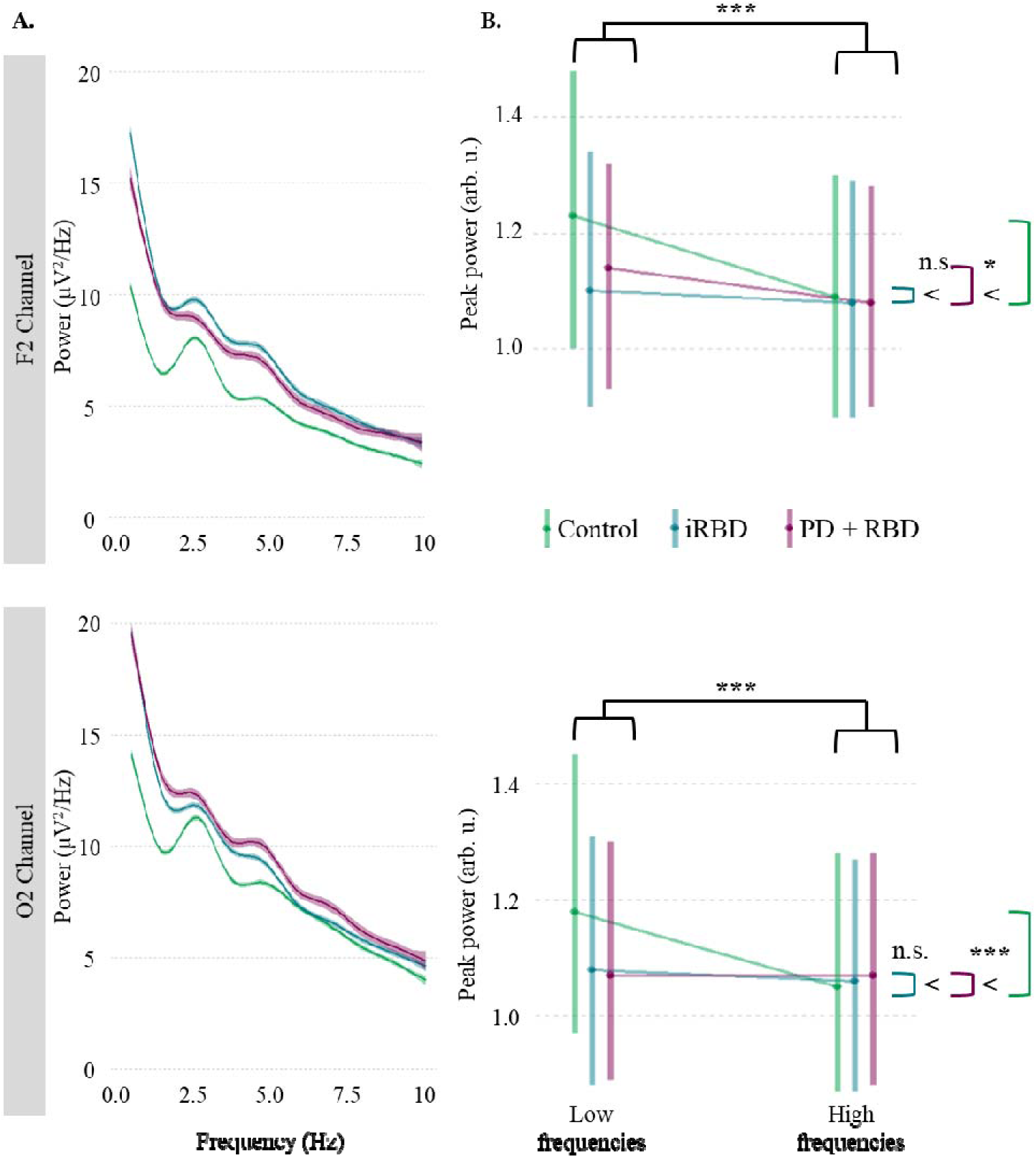
Sawtooth waves (STW) power spectral analysis. A. Generalized additive model (GAM) fits of the power spectrum density of all STW segments separately in each group in F2 (top) and O2 (bottom) channels. Shaded regions represent the 95% confident interval around the fits. A higher peak around 2.5 Hz is observed in controls as compared to the two patient groups. A second peak is observed around 4.5 Hz. B. The power, as reflected by the heights of the peak over and above the aperiodic component, of low frequency peak is greater than the high frequency peak. Further, the imbalance in power between frequency profiles in favor of the low frequencies is stronger for the control participants as compared to the iRBD and PD+RBD ones. Statistics are displayed for each group with the median and the IQR, which are jittered in arbitrary distances on the x-axis within the respective boxplot to increase perceptibility. STW: sawtooth waves; RBD: rapid-eye movement sleep behavior disorder; iRBD: isolated RBD; PD: Parkinson’s disease; IQR: interquartile range, arb. u.: arbitrary units.

The LMM assessing the distribution of STWs segment according to their frequency profiles met the required assumptions after log transformation of the data. The ANOVA revealed a significant main effect of group (Figure 5, F(2,55.50) = 14.46, p < 0.001, η^2^ = 0.25), frequency profile (F(1, 170.63) = 68.08, p < 0.001, η^2^ = 0.60), as well as an interaction between the group and the frequency profile (F(2, 170.63) = 8.67, p < 0.001, η^2^ = 0.15). There was no significant channel main effect (F(1, 170.62) = 0.044, p = 0.83, η^2^ < 0.001). The post hoc analyses revealed that the number of STW segments was higher in the low frequency (< 3.44 Hz; see Table 3 for values) profile as compared to the high frequency profile (t(71.6) = 6.22, p < 0.001). Further, the number of STW segments was lower in the PD + RDB group as compared to the control (t(71.6) = 3.98, p < .001) and the iRBD (t(71.8) = 3.85, p < 0.001) participants. Conversely, the number of STW segments was not significantly different between control and iRBD participants (t(71.6) = 0.45, p = 0.65). Finally the repartition of STW segment numbers with respect to the frequency profile was different between groups with a more imbalance repartition in favor of low frequency in controls than in PD+RBD (t(171) = 4.16, p < 0.001) and iRBD (t(171) = 1.96, p = 0.051) participants. This repartition was also different between iRBD and PD+RBD participants (t(171) = 2.24, p = 0.040). To conclude, these findings showed that the strongest fitted peak had a lower frequency regardless the group. Yet, the imbalance between frequency profiles of STW segments in favor of the low frequencies was stronger for the control and iRBD as compared to the PD+RBD participants.

**Figure 5.**
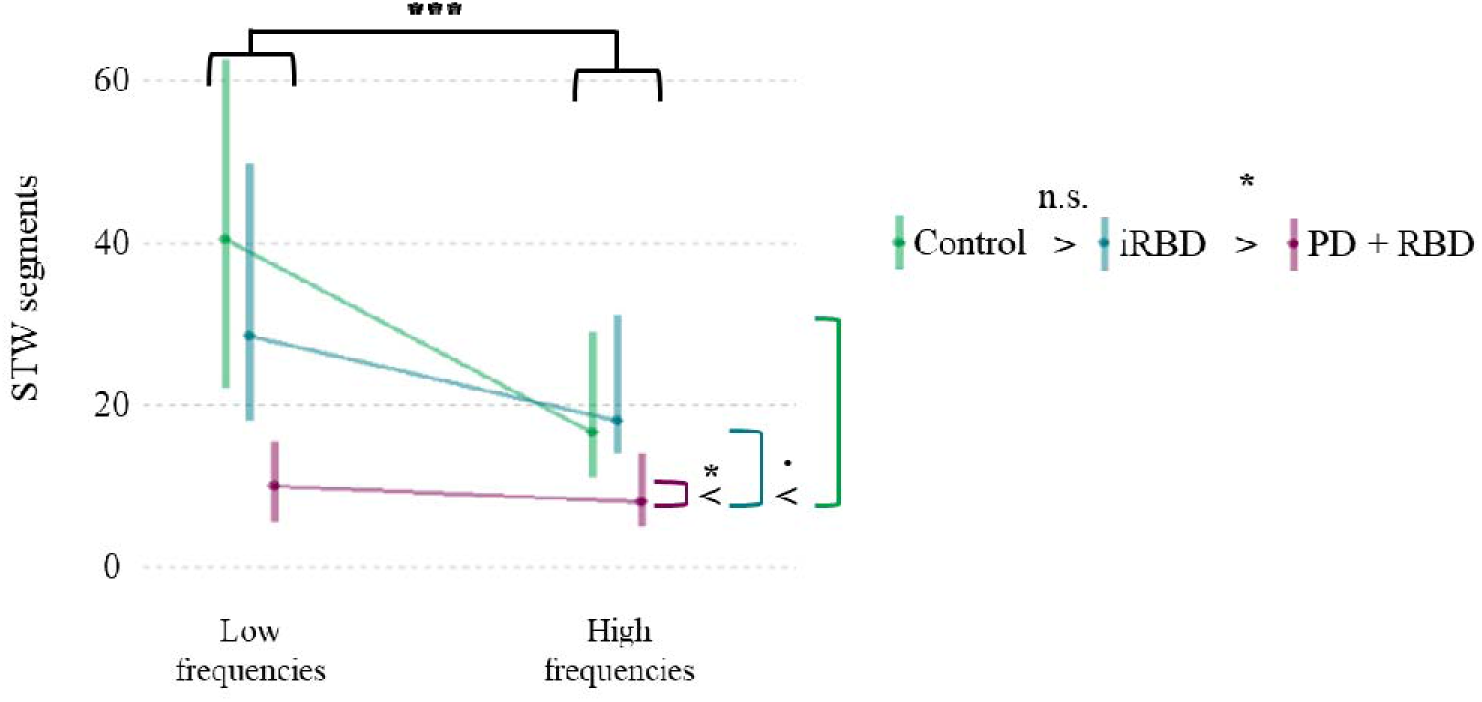
Sawtooth waves (STW) frequency repartition in function of their maximal power in the fits. The stronger fitted peaks have lower frequencies regardless the group. Yet, the imbalance between frequency profiles of STW segments in favor of the low frequencies is stronger for the control and iRBD participants as compared to the PD+RBD ones. Statistics are displayed for each group with the median and the IQR, which are jittered in arbitrary distances on the x-axis within the respective boxplot to increase perceptibility. STW: sawtooth waves; RBD: rapid-eye movement sleep behavior disorder; iRBD: isolated RBD; PD: Parkinson’s disease; IQR: interquartile range.

**Table 3.** Sawtooth waves (STW) segments number and power of peaks per group and per frequency profile. Values are provided as median [IQR]. RBD: REM sleep behavior disorder; iRBD: isolated RBD; PD: Parkinson’s disease.

|  | Control | iRBD | PD+RBD |
| --- | --- | --- | --- |
| Number of segments per participant with maximum peak within each frequency profile |  |  |  |
| Low frequency peak | 40.5 [22.0 – 62.5] | 28.5 [18.0 – 49.7] | 10.0 [5.5 – 15.5] |
| High frequency peak | 16.5 [11.0 – 29.0] | 18.0 [14.0 – 31.0] | 8.0 [5.0 – 14.0] |
| Power of peaks depending on the frequency profile and the topography |  |  |  |
| F2 |  |  |  |
| Low frequency peak | 1.23 $\mu\text{V}^2/\text{Hz}$ [1.00 – 1.48] | 1.10 $\mu\text{V}^2/\text{Hz}$ [0.90 – 1.34] | 1.14 $\mu\text{V}^2/\text{Hz}$ [0.93 – 1.32] |
| High frequency peak | 1.09 $\mu\text{V}^2/\text{Hz}$ [0.88 – 1.30] | 1.08 $\mu\text{V}^2/\text{Hz}$ [0.88 – 1.29] | 1.08 $\mu\text{V}^2/\text{Hz}$ [0.90 – 1.28] |
| O2 |  |  |  |
| Low frequency peak | 1.18 $\mu\text{V}^2/\text{Hz}$ [0.97 – 1.45] | 1.08 $\mu\text{V}^2/\text{Hz}$ [0.88 – 1.31] | 1.07 $\mu\text{V}^2/\text{Hz}$ [0.89 – 1.30] |

| peak |  |  |  |
| --- | --- | --- | --- |
| High frequency peak | 1.05 $\mu\text{V}^2/\text{Hz}$ [0.87 – 1.28] | 1.06 $\mu\text{V}^2/\text{Hz}$ [0.87 – 1.27] | 1.07 $\mu\text{V}^2/\text{Hz}$ [0.88 – 1.28] |

The LMM assessing the power of the STW segment peaks met the required assumptions after square-root transformation of the data. The ANOVA revealed a significant main effect of channel (Figure 4B, F(1, 14157.2) = 18.32, p < 0.001, η^2^ = 0.090), a main effect of frequency profile (F(1, 10961.7) = 82.40, p < 0.001, η^2^ = 0.41), and a trending effect of group (F(2, 57.3) = 2.74, p = 0.073, η^2^ = 0.027). There also was a significant interaction between the group and the frequency profile (F(2, 14223.7) = 48.62, p < 0.001, η^2^ = 0.48).

To investigate these effects further, we performed two separate channel-specific LMM. The post hoc tests consisting of LMMs to assess the power of the STW segment peaks in F2 (see Table 3 for values) revealed a main effect of frequency profile (F(1, 5963.2) = 64.19, p < 0.001, η^2^ = 0.54) and an interaction between group and frequency profile (F(2, 7058.2) = 24.87, p < 0.001, η^2^ = 0.42). There was no main effect of group (F(2,62.4) = 1.94, p = 0.15, η^2^ = 0.033). Further post hoc analyses revealed that the power was higher in the low than in the high frequencies (z = 7.97, p < 0.001) and that this decrease from low to high frequency profiles was significantly greater in the control group as compared to the iRBD (z = 7.01, p < 0.001) and the PD+RBD (z = 3.33, p =0.013) group. Conversely, there was no significant difference between the power decrease from low to high frequencies in the PD+RBD as compared to the iRBD group (z = -1.54, p = 0.12).

In O2 channel, the LMMs assessing the power of the STW segment peaks (see Table 3 for values) revealed a main effect of frequency profile (F(1, 2518.4) = 23.58, p < 0.001, η^2^ = 0.29) and an interaction between group and frequency profile (F(2, 6966.7) = 26.48, p < 0.001, η^2^ = 0.66). There was no main effect of group (F(2,55.94) = 1.73, p = 0.19, η^2^ = 0.043). Further post hoc analyses revealed that the power was higher in the low than in the high frequencies (z = 4.86, p < 0.001) and that this decrease from low to high frequency profiles was significantly greater in the control group as compared to the iRBD (z = 6.19, p < 0.001) and the PD+RBD (z = 5.68, p < 0.001) groups. Conversely, there was no significant difference between the power decrease from low to high frequencies in the PD+RBD as compared to the iRBD group (z = 1.26, p = 0.21). To conclude, the analyses focused on the power in the STW frequency band show that the low frequency peak had a stronger power than the high frequency peak. Further, the imbalance in power between frequency profiles in favor of the low frequencies was stronger for the control participants as compared to the iRBD and PD+RBD ones.

### NREM/REM sleep transitions

A total of 236 REM sleep periods were recorded in the 60 subjects (84 in the controls group, 85 in the iRBD group and 67 in the PD+RBD). Atonia onset could not be marked because of EMG artifact or minimal muscle tone in N3 or N2 stages preceding REM sleep in 52 transitions (21 controls, 14 iRBD and 17 PD+RBD). As a result, the complete sequence of three microstructure features (muscle atonia onset, first STW, first REMs) was analyzed in 186 NREM/REM sleep transitions. The first burst of REMs appeared last in the vast majority of transitions (controls: 63/64, 98.4%; iRBD: 63/71, 88.7%; PD+RBD: 46/51, 90.2%). While atonia onset was first in the majority of transitions in controls group (37/64, 57.8%), STW was first in the majority of transitions for iRBD group (47/71, 66.2%) and PD+RBD group (30/51, 58.8%). The different possible sequences of microstructure features onset during the NREM/REM sleep were not evenly distributed between the three groups (p = 0.0034) (Table 4). Post-hoc analyses revealed that the distribution of the six possible transition sequences differed between controls and iRBD groups (p = 0.00035 (0.001 FDR-corrected)) but not between PD+RBD and control or iRBD groups (p = 0.030 (0.089 FDR-corrected) and p = 0.57 (1.00 FDR-corrected), respectively). These results show that the “atonia-first” triplets are predominant in the control group whereas the “STW-first” sequences are predominant in patients with iRBD and with PD+RBD.

**Table 4.** Sequence of REM sleep microstructure features during NREM/REM sleep transitions. For each group, the number and the percentage of each possible sequence is reported in the table. REM: rapid eye movement; NREM: non-REM; STW: sawtooth waves; RBD: REM sleep behavior disorder; iRBD: isolated RBD; PD: Parkinson’s disease.

| NREM/REM sleep transitions sequence | Controls, n = 64 | iRBD, n = 71 | PD+RBD, n = 51 |
| --- | --- | --- | --- |
| STW - atonia - REMs | 26 (40.6%) | 42 (59.2%) | 28 (54.9%) |
| Atonia - STW - REMs | 37 (57.8%) | 21 (29.6.2%) | 18 (35.3%) |
| STW - REMs - atonia | 0 | 5 (7.0%) | 2 (3.9%) |
| Atonia - REMs - STW | 0 | 3 (4.2%) | 1 (2.0%) |
| REMs - STW - atonia | 0 | 0 | 1 (2.0%) |
| REMs - atonia - STW | 1 (1.7%) | 0 | 1 (2.0%) |

The LMM assessing the NREM/REM sleep transition duration did not meet the required assumptions even after data transformation. We thus performed an ANOVA with permutation test, with group as fixed effect and participant as random effect. This analysis revealed a group main effect (Figure 6, p = 0.0052, η^2^= 0.30). Post-hoc analyses revealed that transition duration was shorter in the control group (119.0 sec [58.5 – 186.1]) as compared to iRBD (212.1 sec [68.5 – 391.4], W = 2650, p = 0.0038 (0.0038 FDR-corrected), r = 0.22) and PD+RBD (375.8 sec [217.6 – 514.6], W = 959, p < 0.001 (< 0.001 FDR-corrected), r = 0.57) groups. Transition duration was shorter in iRBD as compared to PD+RBD (W = 1784, p < 0.001 (< 0.001 FDR-corrected), r = 0.32) participants.

**Figure 6.**
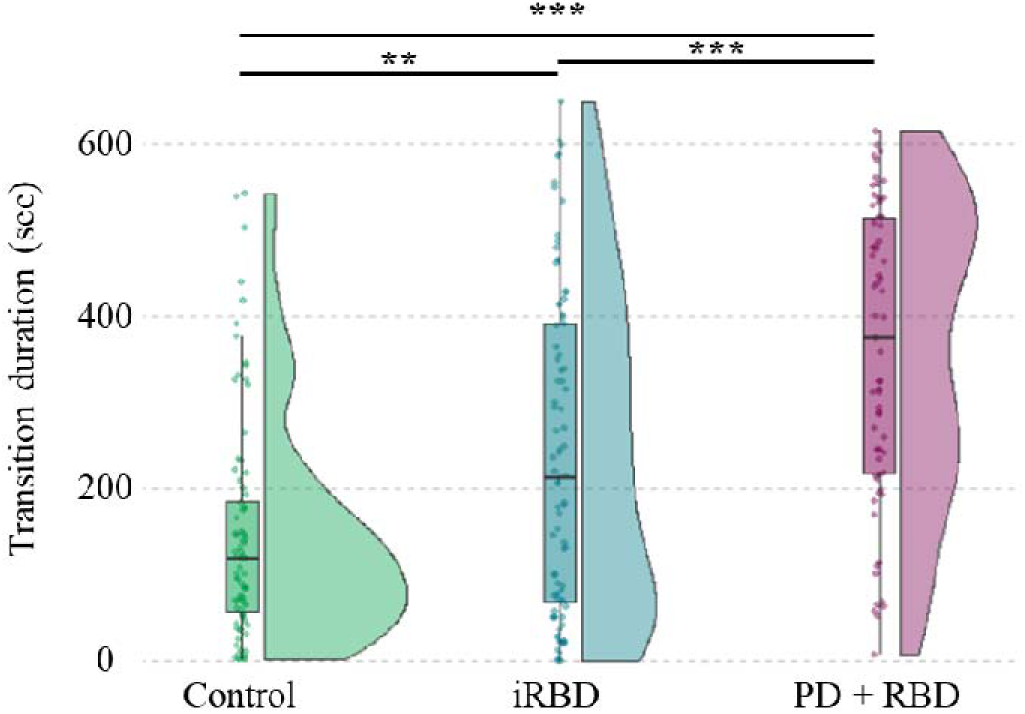
Duration of NREM/REM sleep transitions. The duration of NREM/REM sleep transition (in sec) is longer in the PD+RBD than in the control and iRBD groups. Transitions in the control group are shorter than in the iRBD group. Statistics are displayed for each group with a density plot (kernel density estimate) and a boxplot: the bold line corresponds to the median; lower and upper hinges hinges correspond to first and third quartiles; lower and upper whiskers extend from the lower or upper hinges to the lowest or largest value no further than 1.5xIQR from the hinge. Colored data points correspond to individual values of NREM/REM sleep transitions, jittered in arbitrary distances on the x-axis within the respective boxplot to increase perceptibility. **: p<0.01; ***: p<0.001. REM: rapid eye movement; NREM: non-REM; RBD: REM sleep behavior disorder; iRBD: isolated RBD; PD: Parkinson’s disease; IQR: interquartile range

## DISCUSSION

In this study, leveraging on a systematic analysis of REM sleep microstructure, we observed significant alterations of several features of REM sleep in patients with either iRBD and PD+RBD. In patients with iRBD, the phasic REM sleep ratio was found to be higher than in both controls and patients with PD+RBD. In patients with PD+RBD, we observed alterations in STW bursts, which had lower density and shorter duration than in controls and patients with iRBD. Additionally, both patient groups exhibited STW bursts with distinct spectral characteristics compared to controls with a shift toward higher frequencies. Finally, we identified a gradient in the duration of NREM/REM transitions, with controls having the shortest durations, followed by patients with iRBD, and patients with PD+RBD exhibiting the longest stage transitions.

### Phasic REM sleep is increased in iRBD

While abnormal motor behaviors in RBD appear predominantly during phasic REM sleep, ^[26,27]^ few studies have investigated specifically REM sleep density and phasic REM sleep ratio. In iRBD, data are conflicting: some works have found increased (or a trend toward increased) REM sleep density in patients compared to controls, ^[13,26,28,29]^ while others did not. ^[30–32]^ These discrepancies might be partly attributed to different methods used to quantify REMs and define REM phasic (% time with bursts of at least 3 seconds of REMs) vs REM density (% of 3 sec mini-epochs with versus without a least one REM, or number of REMs / unit of time). They may also result from disparities in samples of RBD patients, with various degrees of disease progression. Indeed, in patients with PD, a decrease in REM density as compared to controls has been reported. ^[56,57]^ Interestingly, REM density is higher in patients with PD+RBD compared to patients with PD without RBD. ^[33]^ With fMRI, it has been shown that REMs bursts are preceded by neuronal activation in the pontine tegmentum, ventroposterior thalamus and primary visual cortex and are time-locked to the recruitment of the putamen and limbic areas. ^[58]^ It can be hypothesized that, as PD progresses, pathological changes in the putamen and cortex may reduce PGO waves, thereby masking the increase in phasic activity associated with RBD. ^[59]^ The mechanisms underlying the increase in REMs observed in RBD remain unknown and may involve either cortical or subcortical processes. REMs are associated with the activation of the limbic cortex, including amygdala, involved in emotional regulation. ^[58]^ In patients with RBD, dream content is altered, with more negative emotions and aggressive themes. ^[60]^ Increased REMs density has been documented in depression ^[61,62]^ and post-traumatic stress disorder ^[63]^ with even RBD-like symptoms in some patients. ^[64]^ The increase in phasic REM sleep revealed here in iRBD might be related to the emotional dysregulation encountered in the patients, associated with nightmares and depressive symptoms. ^[65,66]^ Indeed, RBD is often associated with depression, likely resulting from dysfunction of the limbic circuitry. ^[67,68]^ In addition, restless REM sleep has been suggested to interfere with the overnight resolution of emotional distress, which echoes our findings of increased REM sleep microstructural fragmentation in patients with iRBD. ^[69–71]^ Considering the critical role of REM sleep in emotion regulation, it is possible to speculate on a bidirectional relationship between psycho-behavioral symptoms and REM sleep abnormalities in RBD. Another hypothesis is that α-synuclein-related lesions may contribute to the imbalance between phasic and tonic REM sleep. However, the mechanisms regulating the alternation between phasic and tonic REM sleep remain poorly studied and understood, and pathophysiological conclusions remain speculative. Interestingly, a recent study reported increased REM density in isolated RSWA, suggesting an anatomical or functional overlap between structures regulating muscle tone and REMs. ^[59]^

Finally, one can note that the increase in the proportion of phasic REM sleep over the course of the sleep period was preserved across all groups, with lowest phasic REM sleep quantity in the first quarter of the night. However, patients with PD+RBD displayed an distinct pattern with the highest phasic REM sleep proportion occurring during the second quarter. This altered distribution may reflect an altered homeostatic sleep drive in patients with PD+RBD, ^[72,73]^ underscoring the multifaceted mechanisms of REM sleep dysregulation in this context.

### NREM to REM transitions are lengthened in iRBD and PD+RBD

NREM/REM transitions were longer in patients with iRBD compared to controls and even longer in patients with PD+RBD. This is consistent with previous findings of altered NREM/REM sleep transitions in patients endowed with brainstem damage secondary to bulbar form of post-polio syndrome. ^[45]^ This lengthening may reflect an imbalance between REM-on and REM-off structures, in favor of the latter. In particular, the sublaterodorsal nucleus (SLD, or subcoeruleus nucleus in humans), which is critical for generating muscle atonia in rodents, has been shown to be impaired in RBD. ^[1]^ However, SLD inactivation alone does not suppress REM sleep and cannot fully explain our results. ^[1]^

A potential dysfunction in other structures crucial for REM initiation and maintenance, such as the pedunculopontine nucleus, laterodorsal tegmental nucleus, ventromedial medulla, or thalamic reticular nuclei, might be involved to further explain such prolonged transitions. ^[74,75]^ Alternatively, longer transition durations could result from an hyperactivation of REM-off structures, such as the ventrolateral periaqueductal gray (vlPAG), through damage of inhibitory structures, including the locus coeruleus, the dorsal raphe nucleus, or the lateral hypothalamus. ^[5,76]^ Recently, post-mortem neuropathology in patients with iRBD found that α-synuclein deposits were restricted to various structures of the brainstem (subcoeruleus nucleus, gigantocellular reticular nucleus, laterodorsal tegmentum) and the limbic system (amygdala) while in patients with RBD associated with PD or LBD, the lesions also involved the cortex. ^[77]^ This cortical damage in PD patients may exacerbate the trouble in initiating REM sleep and account for the longest NREM/REM transitions observed in this group. ^[44,78]^ Not only the duration, but also the sequence of events at NREM/REM sleep transitions differed between patients with iRBD or PD+RBD, and controls. Both sequences, STW-first or atonia-first, have been documented in the literature, ^[34,36,45]^ and were also observed here in controls, with a predominance of “atonia-first” triplets. In patients with iRBD and with PD+RBD, “STW-first” sequences were more frequent, suggesting an impairment not only in the maintenance but also in the onset of muscle atonia.

### STW are altered in PD+RBD

STW bursts were fewer in number, shorter, and exhibited altered spectral power, with a reduced predominance of the low-frequency peak in patients with PD+RBD compared to the other groups. The exact generator of STW remains discussed but they have been observed in the thalamus and cortex in human and non-human primates, ^[36,37,43]^ which is corroborated by their alterations in hemispheric stroke and epilepsy. ^[38,39]^ In the iRBD group, the STWs did not show a quantitative but a qualitative difference, with a smaller distinction between spectral profiles in terms of distribution and power. EEG abnormalities have been associated with cognitive impairment ^[8]^ or the risk of phenoconversion ^[10]^ and suggested to reflect a subtle cortical damage. These REM sleep anomalies also parallel those observed for NREM sleep oscillations. In PD, sleep spindles show reduced density, amplitude, and oscillation frequency, along with impaired coupling with slow waves, particularly so when PD is associated with RBD. ^[79–82]^ In iRBD, several studies have reported disruptions in NREM sleep oscillations, including a reduction in (predominantly fast) sleep spindles density ^[82,83]^ and reduced spindle power with impaired slow wave-spindle temporal coupling ^[84]^ suggesting early thalamic and/or cortical dysfunction. Slow waves, including STW, have been suggested to synchronize high-frequency activities across widespread cortical areas during REM sleep,^[36,85]^ as do PGO waves described in animals, ^[86]^ even if their role in cognitive processes including memory consolidation has yet to be demonstrated. Therefore, the functional consequences of impaired STW in PD remain to be further investigated.

### Strengths and Limitations

We have to acknowledge several limitations to our work. First, the number of patients involved was limited, as is often the case for work in this field involving time-consuming visual analysis. ^[26,27]^ In addition, it would have been of interest to include a group of patients with PD without RBD; importantly, the delay between the diagnosis of PD and that of RBD was not known, which could be a source of heterogeneity within this group. The control group consisted of patients referred for sleep apnea screening, who were asymptomatic or paucisymptomatic regarding sleep, as the referrals were primarily made in a cardiovascular context. Importantly, some PD patient were taking medications such as dopaminergic or serotoninergic drugs, which may have influenced REM sleep microstructure. ^[87,88]^ It is also important to note that EEG abnormalities evolve over time in iRBD, ^[14]^ which also introduces heterogeneity when cohorts of patients with varying lengths of RBD progression are studied cross-sectionally. It would be worthwhile monitoring these markers longitudinally. Due to the heterogeneity of our dataset, the number of common electrodes available for spectral analysis was limited, which prevented us from conducting detailed topographical investigations. Finally, while NREM/REM transition duration is fairly easy to estimate in clinical practice, STW analysis is tedious and time-consuming, which underscores the need for developing automatic methods for detecting such graphoelements, as done in spindle analyses. ^[82,83]^

## Conclusion

In RBD, REM sleep disturbances are not limited to muscle tone anomalies. REM sleep microstructure features show alterations, which are easily accessible on the v-PSG performed at diagnosis. They might result from brainstem, subcortical and/or cortical damage with a complex functional outcome. The evolution of these REM sleep microstructural alterations in relation to disease progression and its potential value in predicting the time to and type of phenoconversion in α-synucleinopathy remain to be investigated.

## Data Availability

All data produced in the present study are available upon reasonable request to the authors

## Acknowledgments

JN was supported by the Fondation Pour la Recherche Médicale postdoctoral fellowship (2023-2026), and LC was supported by the Agence Régionale de Santé Auvergne-Rhônes-Alpes master fellowship 2023-2024

